# Typical indicators of neighborhood change used to define gentrification have opposing associations with infant mortality

**DOI:** 10.1101/2024.10.01.24314643

**Authors:** Daria Murosko, Molly Passarella, Diana Montoya-Williams, Roshanak Mehdipanah, Scott Lorch

**Affiliations:** Division of Neonatology, The Children’s Hospital of Philadelphia, Philadelphia, Pennsylvania; Leonard Davis Institute of Health Economics, Philadelphia, Pennsylvania; Department of Pediatrics, Perelman School of Medicine, University of Pennsylvania, Philadelphia, Pennsylvania; Children’s Hospital of Philadelphia PolicyLab, Philadelphia, Pennsylvania; Department of Health Behavior and Health Education, School of Public Health, University of Michigan, Ann Arbor, Michigan

## Abstract

Infant mortality (IM), or death prior to the first birthday, is a key public health metric that increases with neighborhood structural inequities. However, neighborhood exposures shift as communities undergo gentrification, a pattern of neighborhood change defined by increasing affluence (in wealth, education, and housing costs). Gentrification has inconsistent associations with infant health outcomes like IM, which may be due to differing relationships between its composite measures and such outcomes. We designed a retrospective cohort analysis of all births and deaths from 2010-2019 across 4 metropolitan areas in Michigan to determine how gentrification and its neighborhood-change components are associated with risk of IM, using multilevel multivariable logistic regression models. Among 672,432 infants, 0.52% died before 1 year. IM was not associated with gentrification. Census tracts with greater increases in income and education had lower rates of IM, but tracts with greater increases in rent costs had higher rates of IM. In unadjusted models, odds of IM were 40% and 15% lower for infants living in tracts in the top quartile increase in household income and college completion, respectively, compared to infants from tracts with the least amount of change. Odds of IM were also increased 29% in infants from tracts with the most increases in rent, though these differences were attenuated when adjusting for individual social factors. Indicators of increasing community affluence have opposing relationships with IM. Policies and interventions that address rising housing costs may reduce IM.

## Background

Gentrification is a process of neighborhood change characterized by the influx of new, affluent residents into a historically disadvantaged community that results in the elevation of its position relative to a broader urban community.^1^ Gentrification is an increasing phenomenon in urban communities across the US.^2^ Long-term residents in a changing neighborhood are often exposed to erosion of community connectivity, increased housing costs, and fear of displacement, contributing to heightened stress that may ultimately drive adverse health outcomes.^3–5^ Given the unique financial pressures and stresses that occur in early parenting, the health of children in early life may be affected by neighborhood gentrification. Infant mortality (IM), defined as death before the first birthday, represents the extreme of adverse health outcomes, and is a key public health measure of maternal and infant health. IM rates in the United States are greater than other similarly developed nations, and there are wide inequities in IM rates by race and ethnicity, and geographic location. Increased maternal stress has been linked with established drivers of IM such as preterm birth and infant care behaviors.^6,7^ Additionally, maternal mental health conditions are independent risk factors for IM.^8^ However, neighborhood gentrification – and the stress it induces - has not been previously examined as a risk factor for IM.

To advance an understanding of gentrification and health, researchers and policy makers must first operationalize gentrification and identify where neighborhood shifts are occurring. Freeman argued that gentrification is only possible in certain neighborhoods based on their relative deprivation compared to the larger community.^9^ Neighborhoods gentrify if they undergo outsized improvements in sociodemographic factors (often, Census-derived population-level educational attainment) or economic measures (often, housing costs) between two time points. Neighborhoods that don’t exceed this threshold are classified as non-gentrifying. Though alternative definitions of gentrification may set different thresholds of change or include other measures of socioeconomic change (such as share of renters, or accessibility of public transportation),^10^ most continue to conform to Freeman’s original structure of a multi-faceted, composite indicator of neighborhood change. However, there is little agreement on where gentrification is occurring, with minimal overlap in census tracts identified as gentrifying across various methods.^10,11^

The lack of commonality across definitions of gentrification reflects disparate underlying assumptions about the processes driving it.^12^ While Freeman’s original work gives a strong theoretical foundation for which components should be included in the composite index, newer works do not always elucidate an underlying conceptual framework that indicates the hypothesized relationship between the components of the composite measure and the overall construct of gentrification. In addition, local context plays a role; measures of gentrification that have been developed to capture community-specific processes in one region may not be transferrable to other areas.^10^ Thus, a reliance on composite measures obscures the ability to determine the facets of neighborhood change that are most salient to a specific community.

Decomposing the gentrification definitions into their components can address both concerns about causal mechanisms and local context. When singular indicators are compared across many areas, they may identify key, underlying drivers of health outcomes that affect multiple communities. In addition, studying components of gentrification definitions may be important when applying the concept of gentrification to populations different than those in which the index was initially developed. For instance, pregnant people and new parents have unique health needs, such as higher volume of medical appointments, and so may be differentially sensitive to certain components of neighborhood change that exacerbate barriers to healthcare access.

Thus, the primary objective of our analysis was to determine the association between neighborhood gentrification and IM. Secondarily, we decomposed the composite definition of gentrification into its key components and determined the association between these components of neighborhood change and IM. Lastly, we examined these relationships across multiple different metropolitan areas within Michigan, a state with significant variation in current urban revitalization processes, to test the hypothesis that the relationship between gentrification and IM may be heterogeneous even within the same state.

## Methods

### Study Design and Population

In this retrospective cohort analysis, we analyzed linked maternal-infant birth hospitalization, birth certificate, and death certificate data, for all infants born in Michigan from 2010 to 2019, a timeframe chosen to capture the long-term, possibly lagging effects of neighborhood change. Infants were included if they were born at 22 0/7 - 44 6/7 weeks gestational age (GA). We included infants whose birth certificate residence was within 4 of the largest metropolitan areas in the state: Detroit (Wayne, Oakland and Macomb counties), Grand Rapids (Kent, Ionia, Ottawa, Moncaltm and Barry counties), Lansing (Ingham, Eaton and Clinton counties), and Ann Arbor (Washtenaw county). Infants were excluded if birthweight was infeasible for gestational age (>5 standard deviations above or below average for GA), or if residential census tract on birth certificate was missing or could not be linked to census data. The study was designed in accordance with the STrengthening the Reporting of OBservational studies in Epidemiology guidelines and use of these de-identified data was considered non-human subjects research by the institutional review board.^13^

### Exposures of interest

Our primary exposure of interest was census-tract gentrification between 2000 and 2010, calculated using the Ding index of gentrification, which uses Census data.^14^ We obtained 2000 Decennial Census and the 2010 5-year American Community Survey data from Social Explorer, which harmonizes census boundaries across different census tract vintages (accounting for shifts in tract boundaries).^15^ Following the Ding definition, gentrification status was determined using markers of income, educational status and housing costs. Tracts were eligible to be classified as gentrified in this study if they had greater than 50 residents and their median house income (MHI) in 2000 was below the median for their metropolitan area. Tracts with MHI greater than the median in 2000 were designated as “Ineligible” to gentrify. Eligible tracts were designated as “Gentrified” if, between 2000 and 2010, the change in Percent College Completion was above the metro’s median, AND if the 2000-2010 tract change in Median Home Value OR Average Rent was above the metro’s median. Tracts that did not meet both the educational and housing cost criteria were designated as “Did Not Gentrify.”

Our secondary exposures of interests were the components used to calculate the above gentrification index. We investigated tract-specific 2000 to 2010 change in MHI, educational attainment, (percent college completion) and housing costs (both median rent, and median home value). Relative change was measured (difference between 2010 and 2000 values, divided by 2000 value). In addition, we developed quartiles of relative change for each indicator to enhance comparability between metro areas that may have different magnitudes of relative change.

### Outcome of Interest

Our outcome of interest was infant death, defined as death prior to the first birthday.

### Covariate Selection

We included demographic covariates known to be associated with infant death or preterm birth (Table 1). ^16–19^ We also included the metro area and year of birth. Both the effects of gentrification^20,21^ and IM^22^ vary by race and ethnicity, likely secondary to the relationship between structural racism and poverty^23^ and their combined impact on birth and infant health outcomes that increase risk of IM. Thus, using participants self-reported race and ethnicity data from birth certificates, we reported the following categories: Hispanic, Non-Hispanic Asian American/Pacific Islander (AAPI), Non-Hispanic Black (NHB), Non-Hispanic White (NHW), Multiple-Races, and Other Races and Ethnicities. For maternal insurance payor, race and ethnicity, and education level, a missing category indicator was created and incorporated into models.^24^

**Table 1:** Gentrification Status by Metro Area between 2000 – 2010, categorized by Ineligible, Eligible but did not Gentrify (Non-Gent), and Gentrified.

|  | Detroit (n = 450,363) (%) |  |  | Ann Arbor (n = 35,518) (%) |  |  | Lansing (n = 50,206) (%) |  |  | Grand Rapids (n = 136,345) (%) |  |  |
| --- | --- | --- | --- | --- | --- | --- | --- | --- | --- | --- | --- | --- |
| **Gentrification Status** | Ineligible | Non-Gent | Gentrified | Ineligible | Non-Gent | Gentrified | Ineligible | Non-Gent | Gentrified | Ineligible | Non-Gent | Gentrified |
| Observations | 220,486 | 135,373 | 94,504 | 18,048 | 10,888 | 6,582 | 24,300 | 13,593 | 12,313 | 65,311 | 43,073 | 27,961 |
| Maternal Age, years (SD) | 30.2 (5.4)* | 27.0 (5.9) | 27.0 (5.9) | 31.3 (5.3)* | 29.2 (5.6)^ | 28.9 (5.8) | 29.1 (5.4)* | 27.3 (5.7) | 27.3 (5.6) | 29.2 (5.1)* | 27.2 (5.5) | 27.1 (5.6) |
| Maternal Race & Ethnicity |  |  |  |  |  |  |  |  |  |  |  |  |
| Non-Hispanic White | 74.0* | 36.0^ | 38.4 | 75.1* | 57.1^ | 44.0 | 80.5* | 61.2^ | 63.3 | 84.0* | 62.5^ | 62.8 |
| Non-Hispanic Black | 11.9 | 49.6 | 48.3 | 7.0 | 20.2 | 31.2 | 6.2 | 18.6 | 20.4 | 3.0 | 13.1 | 15.4 |
| Hispanic | 4.1 | 8.1 | 7.5 | 5.0 | 6.5 | 6.2 | 5.0 | 9.4 | 9.6 | 8.0 | 18.6 | 16.7 |
| Asian/Pacific Islander | 7.5 | 3.5 | 3.0 | 9.0 | 10.9 | 13.5 | 5.4 | 6.5 | 2.9 | 3.0 | 2.9 | 1.8 |
| Other Race | 0.9 | 1.2 | 1.1 | 1.2 | 1.7 | 1.3 | 1.3 | 2.1 | 1.5 | 0.3 | 0.6 | 0.5 |
| Multiple Races | 1.2 | 1.4 | 1.4 | 2.2 | 2.9 | 3.1 | 1.2 | 1.8 | 1.9 | 1.6 | 2.2 | 2.6 |
| Missing | 0.4 | 0.3 | 0.3 | 0.5 | 0.8 | 0.7 | 0.3 | 0.5 | 0.3 | 0.1 | 0.1 | 0.1 |
| Maternal Education |  |  |  |  |  |  |  |  |  |  |  |  |
| < High School | 0.7* | 3.5^ | 3.9 | 0.4* | 1.1^ | 1.4 | 0.4* | 2.1^ | 1.9 | 1.2* | 5.0^ | 4.7 |
| Some High School | 4.2 | 16.9 | 17.8 | 2.7 | 5.6 | 8.3 | 4.4 | 11.7 | 12.5 | 4.1 | 12.8 | 14.0 |
| High School/GED | 14.6 | 32.9 | 32.5 | 8.6 | 15.5 | 18.0 | 20.8 | 28.9 | 31.3 | 19.3 | 30.2 | 30.3 |
| At least some college | 79.7 | 45.3 | 44.4 | 86.0 | 73.7 | 67.5 | 74.0 | 56.7 | 54.0 | 75.2 | 51.7 | 50.8 |
| Missing | 0.8 | 1.4 | 1.4 | 2.3 | 4.1 | 4.8 | 0.4 | 0.6 | 0.4 | 0.2 | 0.3 | 0.3 |
| Any smoking | 12.2* | 16.9^ | 18.0 | 11.9* | 17.4^ | 18.7 | 17.5* | 23.7^ | 27.0 | 10.9* | 17.1^ | 18.3 |
| Maternal Insurance |  |  |  |  |  |  |  |  |  |  |  |  |
| Private | 74.6* | 37.9^ | 38.9 | 80.2* | 61.0^ | 54.7 | 74.8* | 54.8^ | 52.1 | 77.3* | 50.2^ | 46.7 |
| Government | 24.0 | 61.2 | 60.3 | 16.9 | 36.6 | 43.4 | 24. | 44.5 | 47.2 | 20.9 | 48.0 | 51.6 |
| Self-Pay | 0.7 | 0.6 | 0.5 | 0.6 | 0.5 | 0.6 | 0.6 | 0.5 | 0.6 | 0.5 | 0.5 | 0.6 |
| Other | 0.8 | 0.3 | 0.3 | 2.3 | 1.9 | 1.4 | 0.3 | 0.3 | 0.2 | 1.3 | 1.3 | 1.2 |
| Adequacy of Prenatal Care |  |  |  |  |  |  |  |  |  |  |  |  |
| Inadequate | 6.6* | 16.5 | 16.3 | 8.0* | 12.3^ | 14.6 | 12.4* | 18.6 | 18.6 | 8.8* | 14.3^ | 15.4 |
| Intermediate | 5.9 | 9.6 | 9.6 | 8.4 | 9.1 | 9.5 | 4.9 | 6.1 | 6.2 | 6.2 | 6.4 | 6.5 |
| Adequate | 36.7 | 30.4 | 30.5 | 34.5 | 33.6 | 31.3 | 34.0 | 31.7 | 31.3 | 43.3 | 37.3 | 36.4 |
| Adequate Plus | 44.9 | 33.7 | 33.9 | 44.9 | 40.3 | 39.5 | 47.3 | 42.0 | 42.3 | 39.8 | 39.6 | 39.6 |
| Missing | 5.9 | 9.8 | 9.8 | 4.2 | 4.6 | 5.2 | 1.4 | 1.6 | 1.5 | 1.9 | 2.4 | 2.1 |
| Male Gender at birth | 51.3 | 50.9 | 51.1 | 50.7 | 51.3 | 50.3 | 51.1 | 51.1 | 50.8 | 51.6* | 51.1 | 50.7 |
| Birthweight, kg (IQR) | 3.4* | 3.2 | 3.2 | 3.4* | 3.3^ | 3.3 | 3.4* | 3.3 | 3.3 | 3.4* | 3.3^ | 3.3 |
|  | (3.0, 3.7) | (2.9, 3.5) | (2.9, 3.6) | (3.0-3.7) | (3.0-3.7) | (2.9-3.6) | (3.0-3.7) | (2.9-3.6) | (3.0-3.7) | (3.1-3.7) | (3.0-3.7) | (3.0-3.7) |
| Infant Mortality (per 1000) | 3.9* | 7.9 | 7.4 | 3.5 | 4.0 | 5.0 | 3.7* | 5.1 | 5.3 | 3.5 | 3.9 | 4.2 |

\*p < 0.05 for comparison of Ineligible tracts, Gentrified tracts and Non-Gentrified tracts; ^ p < 0.05 for comparisons of Gentrified vs Non-gentrified tracts. *SD* = standard deviation; *IQR* = interquartile range

### Statistical Analyses

Descriptive statistics were examined across gentrification status (yes, no, ineligible), using chi-squared tests for dichotomous variables and analysis of variance for continuous variables. Given baseline community-level differences and our a priori planned stratified analyses, descriptive statistics were also analyzed by metro area. We conducted mixed-effects logistic regression models of gentrification outcome among eligible tracts and IM with random effects for census tracts nested within metro areas (fixed effects). In adjusted models, we included covariates accounting for metro area, maternal demographics, and year of birth to adjust for temporal trends. Descriptive statistics were also explored for neighborhood change components of gentrification, as described above. We then developed analogous mixed-effects logistic regression models for each component of gentrification (MHI, college attainment, median rent, median home value), exploring the association between relative quartiles of gentrification component change between 2000 and 2010, and IM. As a sensitivity analysis, we tested for an interaction between gentrification status and gentrification component and each of the following covariates: metro area, maternal race and ethnicity, and maternal insurance payor to evaluate whether gentrification differentially affects families by these characteristics. Analyses were also repeated for 2010-2014 only, to determine if there was time-lag variance with the exposure. Statistical analyses were performed using Stata version 18 (StataCorp, College Station, TX).

## Results

Of 1,095,483 eligible births in Michigan between 2010 and 2019, 1,083,205 had census indicators that were linked (98.9%). Of these, 672,432 (62.1%) resided in the metro areas included in the study; 3519 died prior to their first birth (0.52%). Overall, 48.8% lived in neighborhoods that were ineligible to gentrify; 30.2% lived in non-gentrifying neighborhoods and 21.0% lived in neighborhoods that gentrified between 2000 and 2010 (Figure 1; Appendix Table 1). There were significant differences in demographic, pregnancy, and infant characteristics between ineligible, non-gentrifying, and gentrifying neighborhoods. Infants in ineligible neighborhoods had the highest percentage of residents who were NHW, had private insurance, and had completed at least some college. Demographic differences between gentrifying and non-gentrifying neighborhoods varied by metro area (Table 1).

**Figure 1.**
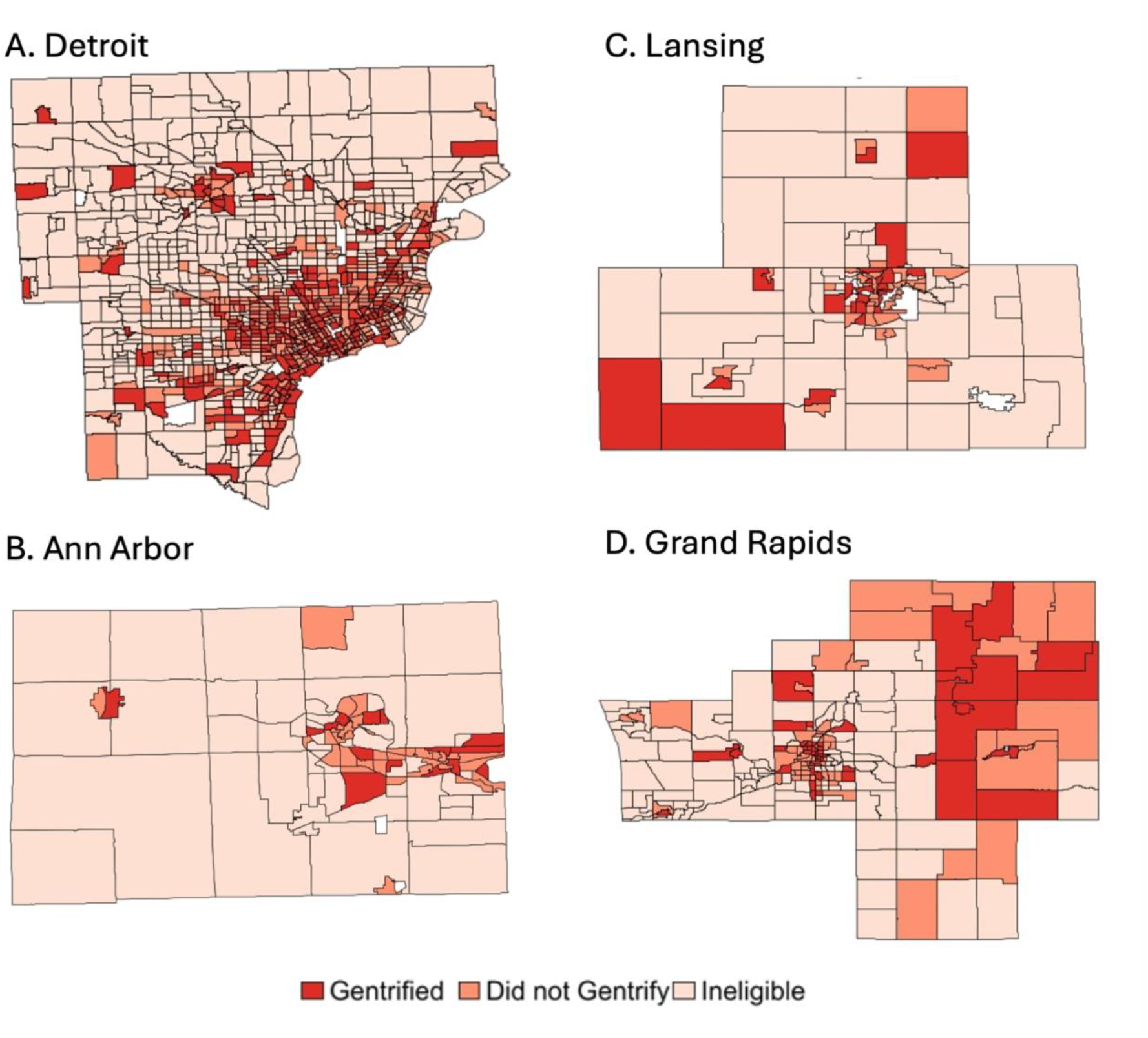
**Census-tract gentrification status by metro area between 2000 and 2010**, using the Ding definition: A) Detroit, B) Ann Arbor, C) Lansing and D) Grand Rapids. Darker red indicates areas that gentrified; lighter red indicates areas that were eligible but did not gentrify; the lightest pink indicates areas that were ineligible to gentrify.

The Ding index of gentrification was not associated with IM in any models (Table 2). The unadjusted odds of IM in a gentrified census tract, compared to a non-gentrified tract was 0.974 (95% CI: 0.875 - 1.084). In adjusted analyses, living in Detroit metro and NHB race were significant risk factors for IM, yet the addition of these covariates had little effect on the estimation of gentrification’s effect. There was no interaction of gentrification status with any of the following: metro area, maternal race and ethnicity, or maternal insurance payor.

**Table 2:** Odds of infant mortality, aOR (95% CI), for birth parents residing in census tracts that gentrified 2000-2010, compared to eligible tracts that did not gentrify^*^.

|  | Unadjusted | Model 1 | Model 2 | Model 3 |
| --- | --- | --- | --- | --- |
| Gentrification | 0.974 (0.875 - 1.084) | 0.975 (0.883 - 1.076) | 0.971 (0.888 - 1.061) | 0.956 (0.876 - 1.043) |
| Metro Area |  |  |  |  |
| Detroit |  | REF | REF | REF |
| Ann Arbor |  | 0.558 (0.433 - 0.721) | 0.677 (0.533 - 0.860) | 0.692 (0.545 - 0.878) |
| Lansing |  | 0.659 (0.539 - 0.805) | 0.824 (0.684 - 0.994) | 0.837 (0.697 - 1.007) |
| Grand Rapids |  | 0.513 (0.444 - 0.593) | 0.669 (0.583 - 0.768) | 0.693 (0.605 - 0.793) |
| Maternal Race & Ethnicity |  |  |  |  |
| Non-Hispanic White |  |  | REFERENCE | REFERENCE |
| Non-Hispanic Black |  |  | 1.942 (1.759 - 2.143) | 1.795 (1.623 - 1.986) |
| Hispanic |  |  | 0.974 (0.815 - 1.165) | 0.884 (0.736 - 1.062) |
| Asian/Pacific Islander |  |  | 0.818 (0.609 - 1.099) | 0.892 (0.663 - 1.199) |
| Other Race |  |  | 1.299 (0.863 - 1.956) | 1.241 (0.822 - 1.873) |
| Multiple Races |  |  | 1.308 (0.930 - 1.839) | 1.216 (0.864 - 1.711) |
| Missing |  |  | 1.812 (0.899 - 3.652) | 1.544 (0.763 - 3.121) |
| Maternal Education |  |  |  |  |
| No high school |  |  |  | REFERENCE |
| Some high school |  |  |  | 1.195 (0.920 - 1.552) |
| High school or GED |  |  |  | 1.010 (0.782 - 1.306) |
| At least some college |  |  |  | 0.788 (0.609 - 1.020) |
| Missing |  |  |  | 2.302 (1.645 - 3.222) |
| Maternal Age |  |  |  |  |
| < 18 years |  |  |  | 1.194 (0.950 - 1.501) |
| 18 – 35 years |  |  |  | REFERENCE |
| > 35 years |  |  |  | 1.132 (0.994 - 1.290) |
| Maternal Insurance |  |  |  |  |
| Private |  |  |  | REFERENCE |
| Government |  |  |  | 1.156 (1.050 - 1.274) |
| Self-Pay |  |  |  | 1.526 (0.940 - 2.476) |
| Other |  |  |  | 0.617 (0.255 - 1.491) |
| Any Smoking |  |  |  | 1.328 (1.200 - 1.471) |

\*Model 3 is also adjusted for year of birth (data not shown)

When gentrification was broken into the 4 components of neighborhood change in the univariate analysis, increasing community income and education levels were associated with lower rates of IM, but the opposite relationship was observed for increasing rent values (Figure 2). Incidence of IM decreased significantly by 28.3% for MHI (p < 0.001), 9.1% for educational attainment (p = 0.006) in tracts below vs above the median change in the change component. Conversely, there was a 28.2% (p < 0.0001) increase in IM between tracts below and above the median change in rent. In other words, in tracts that saw an above average increase in rent costs from 2000 to 2010, frequency of infant death was markedly increased compared to tracts with a below average change in rent. There was no significant change in IM between tracts above and below the median change in home value. When evaluating by metro area, the same trends were seen for Detroit and Ann Arbor metro areas (except in metro Ann Arbor, changes in educational attainment were not significantly associated with IM). In the Lansing and Grand Rapids metro areas, none of the dichotomized neighborhood change indicators were associated with IM.

**Figure 2.**
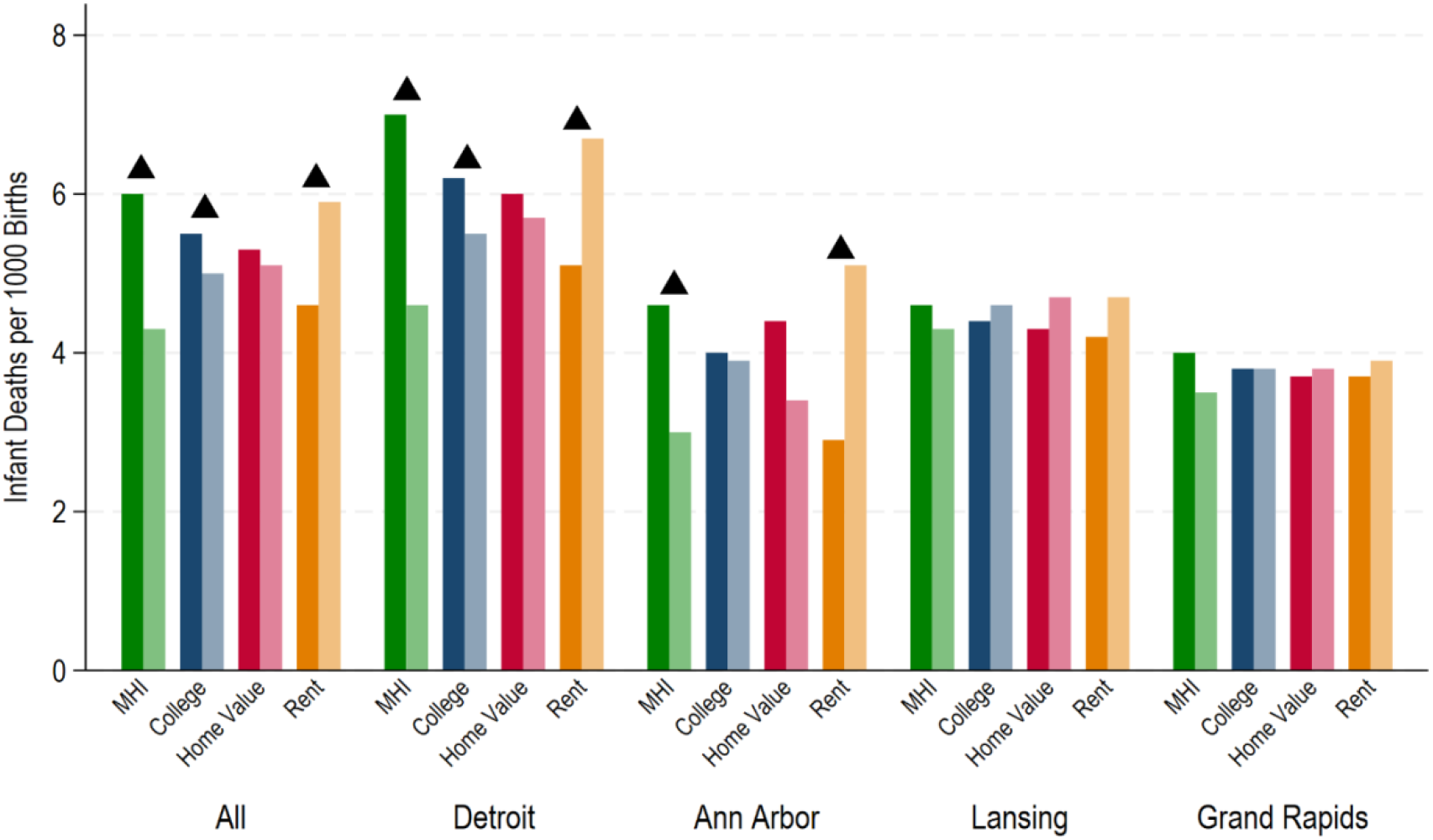
Census-tract rates of infant deaths per 1,000 live births, 2010 - 2019, by neighborhood change components. Neighborhood change components are: 1) Median Household Income (MHI); 2) Proportion college attainment for population > 25 (College); 3) Median Home Value (Homeval); and 4) Median Gross Rent (rent). Darker bars represent relative 2000-2010 change in component that is below the median for the metro area; lighter bars represent relative 2000-2010 change in component that is above the median for the metro area.

In the mixed effects models of neighborhood change indicators and IM, the above patterns persisted (Figure 3, Appendix Table 2). Compared to MHI Quartile 1 (the quartile with the tracts who had the smallest increase in mean household income), infants residing in tracts in MHI Quartile 4 (the greatest increase) had 40% lower odds of IM in unadjusted models (OR 0.60, 95% CI: 0.54 - 0.68). Odds of mortality remained 16% lower in adjusted models (aOR 0.84, 95% CI: 0.76-0.93). The trend was similar for Quartile 1 vs 4 in college attendance change in unadjusted models (OR 0.85, 95% CI: 0.75 - 0.95), but this relationship was not statistically significant when covariates were added. For increases in rent, odds of mortality increased 29% between Quartile 1 and 4 (OR 1.29, 95% CI: 1.14 - 1.46), though this was explained by maternal demographic covariates (aOR 1.01, 95% CI: 0.92 - 1.12). There was no significant association between increasing home value and IM.

**Figure 3.**
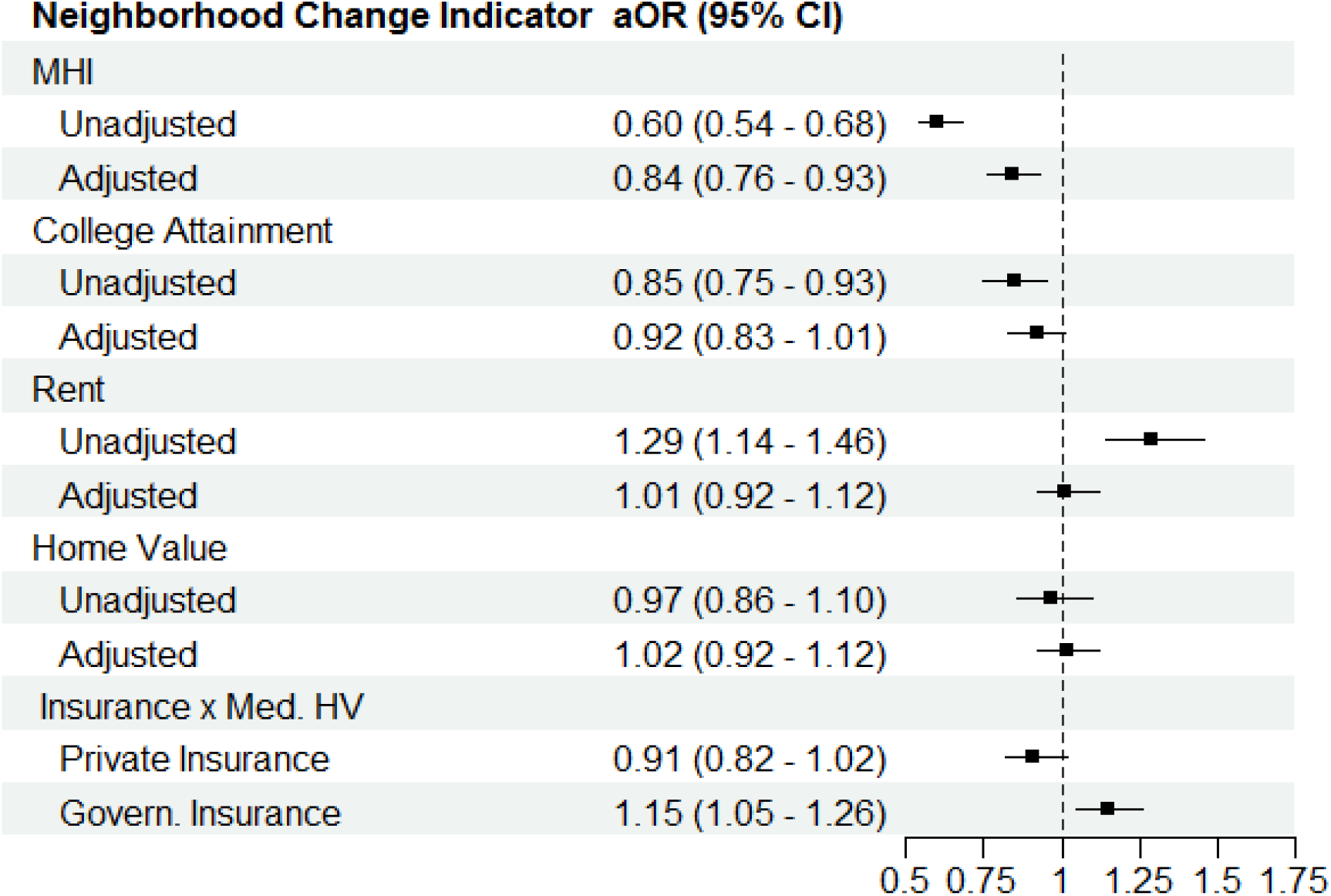
Odds of infant mortality by quartiles of change in gentrification components of neighborhood change. Neighborhood change components are: 1) Median Household Income (MHI); 2) Proportion college attainment for population > 25 years (College Attainment); 3) Median Gross Rent (Rent)l; and 4) Median Home Value (Home Value). Adjusted odds ratios (aORs) and 95% confidence intervals (95% CI) are present for quartile 4, indicating the greatest growth, compared to quartile 1 (least growth) for each neighborhood change components between 2000 and 2010. The interaction between increase in Home Value above the median (Med HV) and insurance (private vs governmental) is also shown. Covariates in adjusted models are metro area, maternal race and ethnicity, age (category), insurance payor, education attainment, and smoking status.

In the sensitivity analyses, there was no significant interaction of any dichotomized neighborhood change component (high vs low change) with metro area or maternal race and ethnicity. Maternal insurance payor (dichotomized as governmental vs private insurance) interacted only with home value change. Living in a census tract with home value that increased above the median was not associated with IM for those on private insurance (aOR 0.91, 95% CI: 0.82, 1.02) (Figure 3; Appendix Table 3). However, infants in those tracts with mothers on governmental insurance at the time of birth had greater risk of IM (aOR 1.15, 95% CI: 1.05, 1.26). When restricted to 2010-2014 births, the same associations between gentrification, neighborhood change components, and IM persisted.

## Discussion

Using an established definition of gentrification, we found no association between gentrification and IM, across multiple metro areas in Michigan. However, when the composite index of gentrification was broken into specific components of neighborhood change, we found that greater increases in neighborhood income and educational attainment predicted lower rates of IM. Conversely, greater increases in rent prices were associated with increased risk of mortality, though this relationship was attenuated when adjusting for maternal socio-demographic covariates.

To our knowledge, the link between neighborhood gentrification and IM has not been previously examined. However, extant analyses have demonstrated conflicting findings on the relationship between gentrification and preterm birth (PTB), a leading cause of IM,^25^ looking across different populations and with different definitions of gentrification.^20,21^ Notably, in both studies found that race was potentially a significant modifier: Huynh found that NHB women experiencing high gentrification were more likely to deliver prematurely, and in Beck’s analysis, gentrification was only protective if looking specifically at Non-White gentrification. In contrast, we found no interaction between gentrification and race and ethnicity across all racial and ethnic groups included in our study population, though dedicated analyses are needed to further assess the relationship between race, gentrification and IM.

Ultimately, analyses focusing on PTB may be poor comparators to our study because multiple pathologies may culminate in infant death. While prematurity contributes significantly to mortality risk, behaviors like sleep positioning, feeding choices and injuries also impact mortality risk, especially over time.^6,25^ While future research could disaggregate the causes of IM to better understand any potential relationship of gentrification with this complex, multifactorial outcome, the trustworthiness of such work will be dependent on the accuracy in which gentrification is defined. We hypothesize that pregnant people and their infants have unique vulnerabilities to specific factors of neighborhood change that are not reflected in the classic, composite definitions of gentrification. Perhaps neighborhood change may have an impact on some pathways to IM but not others.

To address our concern about gentrification measurement, this study decomposed the index of gentrification into separate indicators of neighborhood change, a novel approach to assessing drivers of health. Increasing individual income and educational attainment are established protectors against IM.^26,27^ At the community-level, Collins et al. found that mothers from highly affluent neighborhoods who moved to poorer neighborhoods had higher rates of IM.^28^ Our findings, that neighborhood increases in income and education are associated with lower IM, align with this literature. As we utilized cross-sectional population-aggregated incomes, growth in average income and education may also be driven by in-migration of wealthier, well-educated individuals who, at baseline, have infants with lower risk of IM.^17,19^

We found that increasing neighborhood rent costs was associated with greater risk of IM, a relationship that has not been thoroughly explored in current literature. On a global scale, developed nations with increasing rent costs had lower rates of IM,^29^ though a national evaluation may obscure the hyperlocal variability in the effects of increasing rental costs, particularly when relative increases in rent occur in otherwise impoverished neighborhoods. In addition, the ecological approach prohibits analyses by individual characteristics, such as socioeconomic status (SES) and home ownership status, which are known moderators of the relationship between neighborhood change, housing costs, and health.^30^ For example, U.S.-based analyses have linked increasing housing cost burden to greater risk of pregnancy-related morbidity,^31^ though this effect was more pronounced in populations with lower SES. At baseline, renters are likely to have lower income than homeowners and are more sensitive to rising house costs,^30^ and so they may experience more stress when challenged to absorb this volatility in prices. As our models showed, adjusting for maternal insurance and education – proxies of SES – accounted for the differences that we had found in unadjusted models.

In addition, we found an interaction between home value increases and insurance, with greater IM risk with increasing home value changes only among governmentally-insured individuals. These findings suggest that individuals with low SES (as proxied by government insurance) may be more sensitive to increases in self-reported home values, which often translates into increased monthly mortgages and taxes for homeowners. Aligning with prior findings that low-income homeowners and renters are more likely to be burdened with rising neighborhood housing costs,^32^ our work goes a step further to suggest that this burden may be a risk factor for infant death.

One way in which housing cost burdens might impact health is through their relationship to housing instability, a multi-dimensional construct that captures the spectrum of threats to secure, quality housing,^33^ including evictions, low-quality housing and overcrowding. Rising housing costs are a national concern, as the proportions of American households that were housing cost burdened rose to 23% of homeowners and 50% of renters in 2022,^32^ resulting in greater risk for housing instability leading to evictions and, at its most extreme, homelessness. Among pregnant people, evictions and homelessness have both been linked to adverse birth outcomes including preterm birth and low birthweight.^34,35^ Our analyses linking increased rent costs to IM risk adds a critical new finding to this body of work: addressing the neighborhood housing cost burden among renters may save lives.

Many policy options to curb rising housing cost burdens have been tested at the municipal, state, and federal level. Multi-family zoning amendments and tax incentives for low-income unit construction increase the number of affordable housing units available. Eviction moratoriums and emergency rental assistance authorized by the U.S. Congress, as part of the Coronavirus Aid, Relief and Economic Securities (CARES) Act, helped families stay housed during the economic volatility of the pandemic.^36^ Further work is needed to identify policy choices that provide both an economic benefit and maximize community health. Our work shows that pregnant people and their infants may be a critical subpopulation within which to study such policies.

The effectiveness of such policies may also be dependent on the local context in which they are enacted. As our analysis shows, the relationship between neighborhood change and health varied across the metro areas, which we hypothesize is partially attributable to specific phenotypes of neighborhood change. Between 2000 and 2010, the city of Detroit experienced substantial population loss spurred by lost employment opportunities associated with the automotive industries decentralization, along with another wave of residents moving to the surrounding suburbs.^37^ Like other analyses, we found that gentrification was occurring most in Detroit’s central downtown, consistent with areas of intense revitalization and influx of populations with higher SES.^38^ In contrast, metro Ann Arbor had consistent population growth, paralleling increases in housing costs and income.^15^ These differential processes driving gentrification have differential consequences for health,^12^ as we saw death rates in gentrifying areas of the Detroit metro that were 50% greater than the rates in the Ann Arbor metro area. Nevertheless, significant increases in rent costs were associated with increased IM in both metro areas, suggesting that despite differences in underlying pathways, similar policy levers may be effective to improve health across heterogeneous communities.

The strength of this analysis lies in the large and diverse sample size, particularly as we were able to merge individual data (captured for the entire population of Michigan) to granular, census-tract indicators of neighborhood change. However, there are notable limitations. The scale at which to observe gentrification is frequently a matter of debate: we chose to look at metro areas consisting of multiple counties as done in previous analyses,^39^ but this means we identified gentrifying tracts in suburban areas as well. As a cross-sectional analysis, we are unable to determine how long a mother had been exposed to neighborhood conditions prior to delivery. Specifically, we are unable to determine which mothers may have been displaced from a changing neighborhood prior to delivery; this limitation is common in gentrification analyses.^21,38^ For example, in downtown Detroit where an influx of high SES residents means these areas qualify as gentrified, long-term residents may not have yet been displaced. Previous analyses have shown that the socioeconomic characteristics of displaced people are not substantially different from non-movers,^14^ but longitudinal analyses are needed to determine if there is significant misclassification of exposure in a subset of pregnant people and young children.

## Conclusion

In a large retrospective cohort analysis of multiple metro areas in Michigan, we did not find an association between gentrification and IM. However, when we applied a novel approach of decomposing the composite measure of gentrification into its components, we found that neighborhood improvements in income and education are protective against IM, while increases in neighborhood rent may be harmful. Our results may be used to investigate the impact of policies protecting and promoting affordable housing on infant health, particularly in vulnerable communities. Though it may be impossible to develop interventions that slow the overall process of gentrification, policy experts may be better equipped to match location-dependent changes to targeted interventions around specific components of neighborhood change that are found to be particularly associated with health outcomes in their communities. Further analyses of gentrification should examine aspects of neighborhood change that may be causally related to a health outcome of interest rather than study it solely as a composite exposure, particularly for complex multifactorial health outcomes like IM.

## Data Availability

Data provided by the Michigan Department of Health and Human Resources, who can be contacted with data sharing requests.

