## Appendix Table 1 for "Typical indicators of neighborhood change used to define gentrification have opposing associations with infant mortality"

| Appendix Table 1: Gentrification exposure 2000-2010 and dyad demographics | | | | | |
| --- | --- | --- | --- | --- | --- |
| Eligible tracts | Did not gentrify (n = 202,927) | Gentrified  (n = 141,360) | p-value | Ineligible  (n = 328,145) | p-value* |
| Maternal Age, years | 27.16 (5.78) | 27.15 (5.85) | 0.53 | 29.96 (5.33) | <0.001 |
| Maternal Race/Ethnicity |  |  | <0.001 |  | <0.001 |
| Asian/Pacific Islander | 8,007 (4.0%) | 4,614 (3.3%) |  | 21,499 (6.6%) |  |
| Hispanic | 20,920 (10.3%) | 13,360 (9.5%) |  | 16,402 (5.0%) |  |
| Multiple Races or Ethnicities | 3,379 (1.7%) | 2,497 (1.8%) |  | 4,452 (1.4%) |  |
| Non-Hispanic Black | 77,565 (38.2%) | 54,483 (38.5%) |  | 30,851 (9.4%) |  |
| Non-Hispanic White | 90,156 (44.4%) | 64,569 (45.7%) |  | 251,093 (76.5%) |  |
| Other Race or Ethnicity | 2,315 (1.1%) | 1,490 (1.1%) |  | 2,657 (0.8%) |  |
| Missing | 585 (0.3%) | 347 (0.3%) |  | 1,191 (0.4%) |  |
| Maternal Education |  |  | <0.001 |  | <0.001 |
| No High School | 7,344 (3.6%) | 5,286 (3.7%) |  | 2,507 (0.8%) |  |
| Some High School | 30,532 (15.1%) | 22,818 (16.1%) |  | 13,579 (4.1%) |  |
| High School or GED | 63,131 (31.1%) | 44,236 (31.3%) |  | 51,342 (15.7%) |  |
| At least some College | 99,393 (49.0%) | 67,239 (47.6%) |  | 258,344 (78.7%) |  |
| Missing | 2,527 (1.3%) | 1,781 (1.3%) |  | 2,373 (0.7%) |  |
| Any smoking | 35,376 (17.4%) | 26,668 (18.9%) | <0.001 | 40,332 (12.3%) | <0.001 |
| Maternal Insurance |  |  | <0.001 |  | <0.001 |
| Private | 87,069 (42.9%) | 59,858 (42.3%) |  | 247,628 (75.5%) |  |
| Government | 113,510 (55.9%) | 80,031 (56.6%) |  | 75,506 (23.0%) |  |
| Self-pay | 1,168 (0.6%) | 785 (0.6%) |  | 1,994 (0.6%) |  |
| Other | 1,180 (0.6%) | 685 (0.5%) |  | 3,017 (0.9%) |  |
| Missing | 0 (0.0%) | 1 (0.0%) |  | 0 (0.0%) |  |
| Adequacy of Prenatal Care |  |  | 0.077 |  | <0.001 |
| Inadequate | 32,425 (16.0%) | 22,915 (16.2%) |  | 24,786 (7.6%) |  |
| Intermediate | 17,549 (8.7%) | 12,253 (8.7%) |  | 19,772 (6.0%) |  |
| Adequate | 65,191 (32.1%) | 44,890 (31.8%) |  | 123,548 (37.7%) |  |
| Adequate-Plus | 72,721 (35.8%) | 50,958 (36.1%) |  | 144,668 (44.1%) |  |
| Missing | 15,041 (7.4%) | 10,344 (7.3%) |  | 15,371 (4.7%) |  |
| Infant Gender Male | 103,418 (51.0%) | 72,063 (51.0%) | 0.93 | 168,259 (51.3%) | 0.043 |
| Metro Area |  |  | <0.001 |  | <0.001 |
| Detroit | 135,373 (66.7%) | 94,504 (66.9%) |  | 220,486 (67.2%) |  |
| Ann Arbor | 10,888 (5.4%) | 6,582 (4.7%) |  | 18,048 (5.5%) |  |
| Lansing | 13,593 (6.7%) | 12,313 (8.7%) |  | 24,300 (7.4%) |  |
| Grand Rapids | 43,073 (21.2%) | 27,961 (19.8%) |  | 65,311 (19.9%) |  |
| Gestational Age, weeks | 39 (38-40) | 39 (38-40) | 0.004 | 39 (38-40) | <0.001 |
| Birth Weight, kg | 3.25 (2.90-3.60) | 3.25 (2.89-3.59) | 0.002 | 3.37 (3.02-3.69) | <0.001 |
| Infant Death | 1,355 (0.7%) | 918 (0.7%) | 0.51 | 1,246 (0.4%) | <0.001 |

*p-value for Ineligible vs Gentrified vs Did not gentrify. Mean (SD) presented for maternal age. Median (interquartile range) presented for gestational age and birthweight.
