## Appendix Table 2 for "Typical indicators of neighborhood change used to define gentrification have opposing associations with infant mortality"

| **Appendix 2:** **Full model of odds of infant mortality by quartiles of change in gentrification components of neighborhood change** | | | | | | | | | |
| --- | --- | --- | --- | --- | --- | --- | --- | --- | --- |
| VARIABLES | | Median Household Income | | College Attainment | | Median Gross Rent | | Median Home Value | |
|  |  | Unadjusted | Adjusted | Unadjusted | Adjusted | Unadjusted | Adjusted | Unadjusted | Adjusted |
| Quartile of Change  1^st^ quartile | | REF | REF | REF | REF | REF | REF | REF | REF |
|  | 2nd quartile | 0.76  (0.68 - 0.85) | 0.94  (0.86 - 1.03) | 0.79  (0.70 - 0.88) | 1.00  (0.90 - 1.10) | 1.05  (0.93 - 1.18) | 0.93  (0.84 - 1.03) | 0.86  (0.77 - 0.97) | 1.00  (0.91 - 1.10) |
|  | 3rd quartile | 0.66  (0.59 - 0.74) | 0.89  (0.80 - 0.98) | 0.77  (0.68 - 0.86) | 0.97  (0.88 - 1.07) | 1.34  (1.19 - 1.51) | 1.03  (0.93 - 1.14) | 0.85  (0.75 - 0.95) | 1.05  (0.95 - 1.15) |
|  | 4th quartile | 0.60  (0.54 - 0.68) | 0.84  (0.76 - 0.93) | 0.85  (0.75 - 0.95) | 0.92  (0.83 - 1.01) | 1.29  (1.14 - 1.46) | 1.01  (0.92 - 1.12) | 0.97  (0.86 - 1.10) | 1.02  (0.92 - 1.12) |
| Metro Area  Detroit | |  | REF |  | REF |  | REF |  | REF |
|  | Ann Arbor |  | 0.78  (0.65 - 0.93) |  | 0.79  (0.66 - 0.95) |  | 0.80  (0.67 - 0.95) |  | 0.80  (0.67 - 0.95) |
|  | Lansing |  | 0.91  (0.79 - 1.04) |  | 0.91  (0.79 - 1.05) |  | 0.91  (0.79 - 1.04) |  | 0.90  (0.78 - 1.04) |
|  | Grand Rapids |  | 0.80  (0.73 - 0.89) |  | 0.81  (0.73 - 0.90) |  | 0.81  (0.74 - 0.90) |  | 0.81  (0.73 - 0.90) |
| Education  No high school | |  | REF |  | REF |  | REF |  | REF |
|  | Some high school |  | 1.24  (0.98 - 1.59) |  | 1.24  (0.97 - 1.58) |  | 1.24  (0.97 - 1.58 |  | 1.23  (0.96 - 1.57) |
|  | High school diploma/GED |  | 1.05  (0.83 - 1.34) |  | 1.04  (0.82 - 1.32) |  | 1.05  (0.83 - 1.33) |  | 1.04  (0.82 - 1.32) |
|  | Some college |  | 0.82  (0.64 - 1.04) |  | 0.80  (0.63 - 1.02) |  | 0.80  (0.63 - 1.02) |  | 0.80  (0.63 - 1.01) |
|  | Missing |  | 2.71  (2.01 - 3.64) |  | 2.70  (2.01 - 3.64) |  | 2.70  (2.01 - 3.64) |  | 2.66  (1.97 - 3.58) |
| Maternal Age  18-35 years | |  | REF |  | REF |  | REF |  | REF |
|  | < 18 years |  | 1.13  (0.92 - 1.40) |  | 1.13  (0.91 - 1.40) |  | 1.13  (0.92 - 1.40) |  | 1.13  (0.92 - 1.40) |
|  | > 35 years |  | 1.13  (1.03 - 1.25) |  | 1.12  (1.02 - 1.23) |  | 1.13  (1.02 - 1.24) |  | 1.13  (1.02 - 1.24) |
| Maternal Insurance  Private insurance | |  | REF |  | REF |  | REF |  | REF |
|  | Public Insurance |  | 1.25  (1.15 - 1.35) |  | 1.26  (1.17 - 1.36) |  | 1.26  (1.17 - 1.37) |  | 1.27  (1.17 - 1.37) |
|  | Self-Pay |  | 1.51  (1.03 - 2.22) |  | 1.52  (1.04 - 2.23) |  | 1.54  (1.05 - 2.26) |  | 1.53  (1.04 - 2.24) |
|  | Other |  | 0.63  (0.35 - 1.15) |  | 0.63  (0.35 - 1.15) |  | 0.65  (0.36 - 1.17) |  | 0.63  (0.35 - 1.15) |
| Any smoking | |  | 1.30  (1.19 - 1.41) |  | 1.30  (1.19 - 1.42) |  | 1.29  (1.19 - 1.41) |  | 1.29  (1.19 - 1.41) |
| Race and Ethnicity | |  |  |  |  |  |  |  |  |
|  | Asian American/Pacific Islander |  | 0.81  (0.66 - 0.99) |  | 0.81  (0.66 - 0.99) |  | 0.82  (0.67 - 1.00) |  | 0.80  (0.65 - 0.99) |
|  | Hispanic |  | 0.94  (0.81 - 1.09) |  | 0.95  (0.82 - 1.10) |  | 0.94  (0.81 - 1.09) |  | 0.95  (0.82 - 1.11) |
|  | Non-Hispanic Black |  | 1.89  (1.74 - 2.05) |  | 1.94  (1.79 - 2.11) |  | 1.93  (1.78 - 2.09) |  | 1.96  (1.81 - 2.13) |
|  | Non-Hispanic White |  | REF |  | REF |  | REF |  | REF |
|  | Multiple races |  | 1.43  (1.11 - 1.84) |  | 1.44  (1.12 - 1.86) |  | 1.43  (1.11 - 1.84) |  | 1.43  (1.11 - 1.84) |
|  | Other race |  | 1.31  (0.94 - 1.82) |  | 1.32  (0.95 - 1.84) |  | 1.34  (0.96 - 1.86) |  | 1.31  (0.94 - 1.83) |
|  | Missing Race |  | 0.94  (0.50 - 1.75) |  | 0.94  (0.50 - 1.76) |  | 0.95  (0.51 - 1.79) |  | 0.94  (0.50 - 1.77) |
|  | Quartiles of change indicate increasing growth: 1^st^ quartiles grew the smallest relative amount between 2000 and 2010; 4^th^ quartiles had the largest amount of growth over the same time perio | | | | | | | | |
