## Appendix Table 3 for "Typical indicators of neighborhood change used to define gentrification have opposing associations with infant mortality"

| Appendix 3: Maternal and infant demographics by change in neighborhood change indicator, dichotomized to high and low change, % | | | | | | | | | | | | |
| --- | --- | --- | --- | --- | --- | --- | --- | --- | --- | --- | --- | --- |
|  | Median Household Income | | | College Attainment | | | Median Gross Rent | | | Median Home Value | | |
|  | Low Change | High Change | p-value | Low Change | High Change | p-value | Low Change | High Change | p-value | Low Change | High Change | p-value |
| Metro Area |  |  | <0.001 |  |  | <0.001 |  |  | <0.001 |  |  | <0.001 |
| Detroit | 65.6 | 68.6 |  | 66.8 | 67.2 |  | 66.1 | 67.9 |  | 66.4 | 67.7 |  |
| Ann Arbor | 5.9 | 4.6 |  | 4.9 | 5.7 |  | 5.4 | 5.2 |  | 5.2 | 5.4 |  |
| Lansing | 6.8 | 8.2 |  | 6.8 | 8.1 |  | 7.4 | 7.6 |  | 7.2 | 7.8 |  |
| Grand Rapids | 21.8 | 18.6 |  | 21.5 | 19.1 |  | 21.1 | 19.4 |  | 21.3 | 19.2 |  |
| Premature birth | 10.7 | 9.7 | <0.001 | 10.2 | 10.3 | 0.71 | 9.8 | 10.8 | <0.001 | 10.3 | 10.2 | 0.64 |
| Birthweight, grams* | 3280  (2920-3615) | 3340  (2990-3668) | <0.001 | 3295  (2948-3630) | 3317  (2960-3651) | <0.001 | 3320  (2977-3657) | 3289  (2928-3626) | <0.001 | 3290  (2948-3629) | 3317  (2965-3656) | <0.001 |
| Maternal Age |  |  | <0.001 |  |  | <0.001 |  |  | <0.001 |  |  | <0.001 |
| < 18 years | 1.9 | 1.1 |  | 1.6 | 1.5 |  | 1.1 | 2.0 |  | 1.6 | 1.5 |  |
| 18 - 35 years | 84.6 | 81.1 |  | 82.9 | 83.1 |  | 82.8 | 83.2 |  | 84.3 | 81.6 |  |
| > 35 years | 13.5 | 17.8 |  | 15.5 | 15.5 |  | 16.1 | 14.8 |  | 14.2 | 16.9 |  |
| Maternal Race/Ethnicity |  |  | <0.001 |  |  | <0.001 |  |  | <0.001 |  |  | <0.001 |
| Asian/PI | 4.8 | 5.4 |  | 5.7 | 4.5 |  | 6.6 | 3.4 |  | 5.0 | 5.1 |  |
| Hispanic | 8.5 | 6.4 |  | 8.2 | 6.9 |  | 7.0 | 8.1 |  | 6.9 | 8.2 |  |
| Multiple Races | 1.0 | 0.9 |  | 1.0 | 0.9 |  | 1.0 | 0.9 |  | 1.2 | 0.7 |  |
| Non-Hispanic Black | 31.7 | 15.8 |  | 26.6 | 22.0 |  | 18.2 | 30.8 |  | 26.8 | 21.5 |  |
| Non-Hispanic White | 52.0 | 69.8 |  | 56.8 | 63.8 |  | 65.2 | 55.0 |  | 58.2 | 62.7 |  |
| Other Race | 1.7 | 1.4 |  | 1.5 | 1.6 |  | 1.6 | 1.5 |  | 1.6 | 1.4 |  |
| Missing | 0.3 | 0.3 |  | 0.3 | 0.3 |  | 0.4 | 0.3 |  | 0.3 | 0.3 |  |
| Maternal Education |  |  | <0.001 |  |  | <0.001 |  |  | <0.001 |  |  | <0.001 |
| No high school | 2.8 | 1.6 |  | 2.4 | 2.1 |  | 1.8 | 2.8 |  | 2.4 | 2.1 |  |
| Some high school | 12.3 | 7.3 |  | 10.2 | 9.8 |  | 7.7 | 12.5 |  | 10.3 | 9.6 |  |
| High School/GED | 27.3 | 19.4 |  | 23.7 | 23.5 |  | 21.3 | 26.1 |  | 25.7 | 21.4 |  |
| At least some college | 56.4 | 70.9 |  | 62.7 | 63.7 |  | 68.4 | 57.6 |  | 60.5 | 66.1 |  |
| Missing | 1.2 | 0.8 |  | 1.0 | 1.0 |  | 0.9 | 1.1 |  | 1.2 | 0.8 |  |
| Smoking in Pregnancy | 16.3 | 14.0 | <0.001 | 14.4 | 16.0 | <0.001 | 14.3 | 16.2 | <0.001 | 15.6 | 14.9 | <0.001 |
| Maternal Insurance |  |  | <0.001 |  |  | <0.001 |  |  | <0.001 |  |  | <0.001 |
| Private | 51.2 | 67.2 |  | 57.8 | 59.5 |  | 63.7 | 53.1 |  | 55.7 | 61.9 |  |
| Government | 47.6 | 31.4 |  | 40.9 | 39.2 |  | 34.8 | 45.7 |  | 43.0 | 36.7 |  |
| Self-Pay | 0.6 | 0.6 |  | 0.6 | 0.6 |  | 0.6 | 0.6 |  | 0.6 | 0.5 |  |
| Other Insurance | 0.7 | 0.8 |  | 0.7 | 0.7 |  | 0.9 | 0.6 |  | 0.7 | 0.8 |  |
| Infant Death | 0.6 | 0.4 | <0.001 | 0.6 | 0.5 | 0.006 | 0.5 | 0.6 | <0.001 | 0.5 | 0.5 | 0.26 |

*Birthweight is presented as median (interquartile range). All other indicators as %.
